# Development of the NeuroFlow Severity Score and Comparison With Validated Measures for Depression and Anxiety

**DOI:** 10.1101/2021.02.04.21251158

**Authors:** William Lynch, Michael L. Platt, Adam Pardes

## Abstract

**Purpose:** Although depression and anxiety are the leading causes of disability in the United States, respectively, fewer than half of people diagnosed with these conditions receive appropriate treatment, and fewer than 10% receive measurement-based care (MBC), which is defined as behavioral health care based on and adapted in response to patient outcomes data collected throughout treatment. The NeuroFlow platform was developed with the goal of making MBC easier to deliver and more accessible within integrated behavioral health care. Data from over 3,000 users of the NeuroFlow platform were used to develop the NeuroFlow Severity Score (NFSS), a potential new measure for depression and anxiety. To begin evaluating the potential usefulness of this new measure, NFSSs were compared with validated measures for depression and anxiety, the Personal Health Questionnaire-9 (PHQ-9) and Generalized Anxiety Disorder-7 (GAD-7) scale, and clinician assessment.

**Methods:** The NFSS platform is used to record patient-reported and passively collected data related to behavioral health. An artificial-intelligence derived algorithm was developed that condenses this large number of measurements into a single score for longitudinal tracking of an individual’s depression and anxiety symptoms. Linear regression and Bland-Altman analyses were used to evaluate relationships and differences between NFSS and PHQ-9 or GAD-7 scores from over 35,000 NeuroFlow users. The NFSS was also compared to assessment by a panel of expert clinicians for a subset of 250 individuals.

**Results:** Linear regression results showed a strong correlation between NFSS and PHQ-9 (r=.74, P<.001) and GAD-7 (r=.80, *P*<.001) changes. There was also a strong positive correlation between the NFSS and expert panel clinical assessment (r=.80-.84, P<.001). Bland-Altman analysis and evaluation of outliers on regression analysis, however, show that the NFSS has significant differences from the PHQ-9.

**Conclusions:** Clinicians can reliably use the NFSS as a proxy measure for monitoring symptoms of depression and anxiety longitudinally. The NFSS may identify at-risk individuals who are not identified by the PHQ-9. Further research is warranted to evaluate the sensitivity and specificity of the NFSS.

## Background

### Accessing Behavioral Health Care

In the United States, 1 in 5 adults experience mental illness each year, and suicide is the 10th leading cause of death.^1,2^ Anxiety and depression are leading causes of disability in the US, costing over $300 billion annually in care and lost productivity.^1-3^ Both depression and anxiety correlate with suicide attempts, which are increasing in the United States; in 2018, there were 1.4 million suicide attempts, and 1 in 3 caused death (48,344 deaths).^4^

Despite the magnitude of the problem, fewer than half of people affected by anxiety and depression receive appropriate treatment. The stigma of behavioral health disorders and the symptoms themselves (eg, apathy, fear, isolation) factor into the 23% of people who do not seek care.^6^ Access to care, however, accounts for a much larger proportion of undertreatment and is limited by out-of-pocket costs, unequal geographic distribution of providers, and an overall shortage of providers.^7-9^

### Integrated Measurement-Based Behavioral Health Care

Measurement-based care (MBC) can be broadly defined as the use of continuous monitoring of patient data to inform and, as needed, redirect clinical care.^10^ In the field of behavioral health, MBC is considered an evidence-based practice, which is defined as relying on the use of modern data from controlled scientific studies reported in the published literature to make reasonable and conscientious decisions about clinical care.^11^ As a framework for behavioral health care, MBC uses validated clinical scales to measure symptoms of anxiety and depression before or during a patient-clinician interaction, and then uses those measurements to guide further clinical interventions.^10^ Despite strong evidence in the published literature that MBC improves behavioral health outcomes,^10,12-15^ fewer than 20% of behavioral health care providers use MBC.^14,15^ Research has shown that barriers to use of MBC include, but are not limited to, the costs in time and resources to take measurements and concerns about potential violations of confidentiality with paper and pen measurements.^13,14^

Integrating behavioral health care into primary care has been proposed as a means of improving access to behavioral health care and is supported by both the American Association of Family Physicians and the American Psychiatric Association.^16,17^ In the integrated behavioral health model, the primary care clinician screens patients for symptoms of mental health disorders at every visit and recommends and prescribes further assessment and treatment for behavioral health conditions (eg, depression or anxiety) as needed. Patients are followed by a care manager, and a psychiatrist is available to consult with the primary care clinician as needed for any cases in which management is not straightforward. This model could help address the shortage of behavioral health care providers and low levels of access to behavioral health care, considering that data from 2015 showed approximately 75% of people in the US had a primary care provider (although that was gradually decreasing).^18^ Integrated behavioral health may also help address the unequal distribution of behavioral health care providers because in this model, the behavioral health care professional available for consultation need not be physically in the same location as the primary care provider. This model, however, is not yet widely used. with only 4.3% of primary care visits in 2016 including screening for behavioral health symptoms according to the 2016 National Ambulatory Medical Care Survey.^19^ With such low levels of depression screening during primary care visits, it is perhaps not surprising that data are limited regarding use of MBC within integrated behavioral health care. It is known, however, that time, resources, and confidentiality are obstacles for use of MBC by behavioral health providers who typically have 45 minutes with a patient weekly or biweekly.^13,14^ In that context, it is easy to imagine that it would be difficult for primary care clinicians to make use of MBC.

There is also a paucity of data regarding how much time primary care physicians spend with individual patients. Typically, visits are scheduled every 15 minutes, and data suggest the average visit may last 17 to 21 minutes,^20,21^ although a time-and-motion study suggests only 53% of that time (9-11 minutes) is spent engaging with the patient directly.^22^ Although current validated measures of depression and anxiety, the Personal Health Questionnaire-9 (PHQ-9) and the Generalized Anxiety Disorders-7 (GAD-7), are brief and take less than 5 minutes to administer, this still means providing MBC could take 10% to 30% of a visit and 20% to 60% of severely limited face-to-face to time, during which many other clinical tasks must also be performed.

### Behavioral Health Care Needs Remain

In summary, whether because of access to care, short times spent with physicians, or shortages of behavioral health clinicians, it is clear that many individuals with depression and anxiety are not receiving integrated behavioral health care, much less MBC, despite the evidence that it improves outcomes. Considering the epidemic proportions of depression and anxiety and related disability and mortality, there is an urgent need to find novel ways to make it easier for primary care clinicians to provide MBC in an integrated behavioral health care model. The NeuroFlow platform was built with that goal of improving access to MBC at scale in an integrated behavioral health care model by having assessments, including but not limited to the PHQ-9 and GAD-7, delivered to and completed by the patient outside of the clinical visit in a manner that fits seamlessly into the patient’s day-to-day life instead of adding to a list of tasks that is already more than can be done in many clinical visits. Clinicians with access to the NeuroFlow platform send individualized links to their patients, which allows the patient to download the NeuroFlow mobile app onto their own smartphone or access NeuroFlow through an internet site. The app and platform are HIPAA-compliant, assure confidentiality, and fit easily into patients’ daily activities. Through the NeuroFlow app or desktop interface, patients record, track, and report their mood, sleep quality, and stress level, and complete PHQ-9 and GAD-7 scales regularly. The app also provides engaging educational videos based on users’ self-reported scores and symptom changes over time. Although the latter features are not necessarily interventional or measurement oriented, these serve a goal of maintaining user engagement and lasting behavior change over time by supporting users in taking charge of their own health. Clinicians access patient-reported measures through an internet-based clinician dashboard that integrates directly into electronic health record (EHR) systems to keep track of individual patient’s measurements and receive reports of their patients’ scores. This ensures that taking the measurements required for MBC does not further reduce the already small amount of time clinicians have with patients. Instead of performing screening assessments, they can immediately see which of their patients may require a behavioral health intervention.

Notably, there are some common concerns regarding smartphone apps for behavioral health care, Among these are that digital measurements are not necessarily evidence-based, may not provide treatment that is equivalent to that received from a trained clinician, and often have a low frequency of use and engagement by patients. Often the measures used in digital applications were validated as paper and pen(cil) instruments, and it is not clear if digital versions have the same sensitivity and specificity. The number of available behavioral health applications for smartphones is in the tens of thousands,^23^ whereas a search of Pubmed.gov for clinical studies of such apps returns results in the low thousands. The applications also deliver a wide variety of measures and interventions, making comparisons among apps and between apps and human-delivered (in-person or via telehealth) clinical interventions difficult, if not impossible, to make without such systematic studies. Concerns about persistence of any effects are related to the fact that use of behavioral health apps on average falls off precipitously with only 3.3% of downloaded behavioral-health apps used for more than 30 days.^24^ In response to these important concerns, we note that the 30-day retention rate for NeuroFlow users to date is 70%, as the platform seeks to aid rather than replace clinicians. In addition, as part of our efforts to continue simplifying and, thereby, increasing adoption of MBC by clinicians and patients, we developed the NeuroFlow Severity Score (NFSS)—a single composite, non-diagnostic measure of anxiety and depression that can be tracked over time and have begun the research to validate this as an evidence-based measure. Herein we report data comparing the NFSS with the Personal Health Questionnaire-9 (PHQ-9), the Generalized Anxiety Disorder-7 (GAD-7) scale, and to expert clinician consensus.

## Methods

### Development of the NFSS

To create the composite NFSS, we developed a proprietary algorithm that utilizes measures recorded as patients use the NeuroFlow platform. The dataset included de-identified records of over 3,000 people who used the NeuroFlow platform after it was assigned to them by a clinician between 2018 and 2019. Variables include scores on PHQ-9 or GAD-7 measures taken within the app, which are 27- and 21-point scales, respectively; self-reported sleep-quality measures, using a scale of 0-10; self-reported mood measures on a scale of 0-10, active behavioral health treatment (yes/no), frequency of specific activities within the app (collected passively), and whether the individual had endorsed having suicidal ideation (yes/no). For each measure that factors into the NFSS, descriptive statistics were calculated to determine the distribution of variables; central tendencies through mean and median; and the spread of variable values through range, standard deviation, and variance. Variables were assigned positive or negative weights based on expert clinician input, and these were combined, using a proprietary artificial intelligence algorithm, to produce an NFSS score that ranges from 1 to 5, with 1 reflecting a low risk for depression and anxiety and 5 reflecting a high risk for depression and anxiety. See Figure 1 for an overview of how the NFSS was created and evaluated.

**Figure 1.**
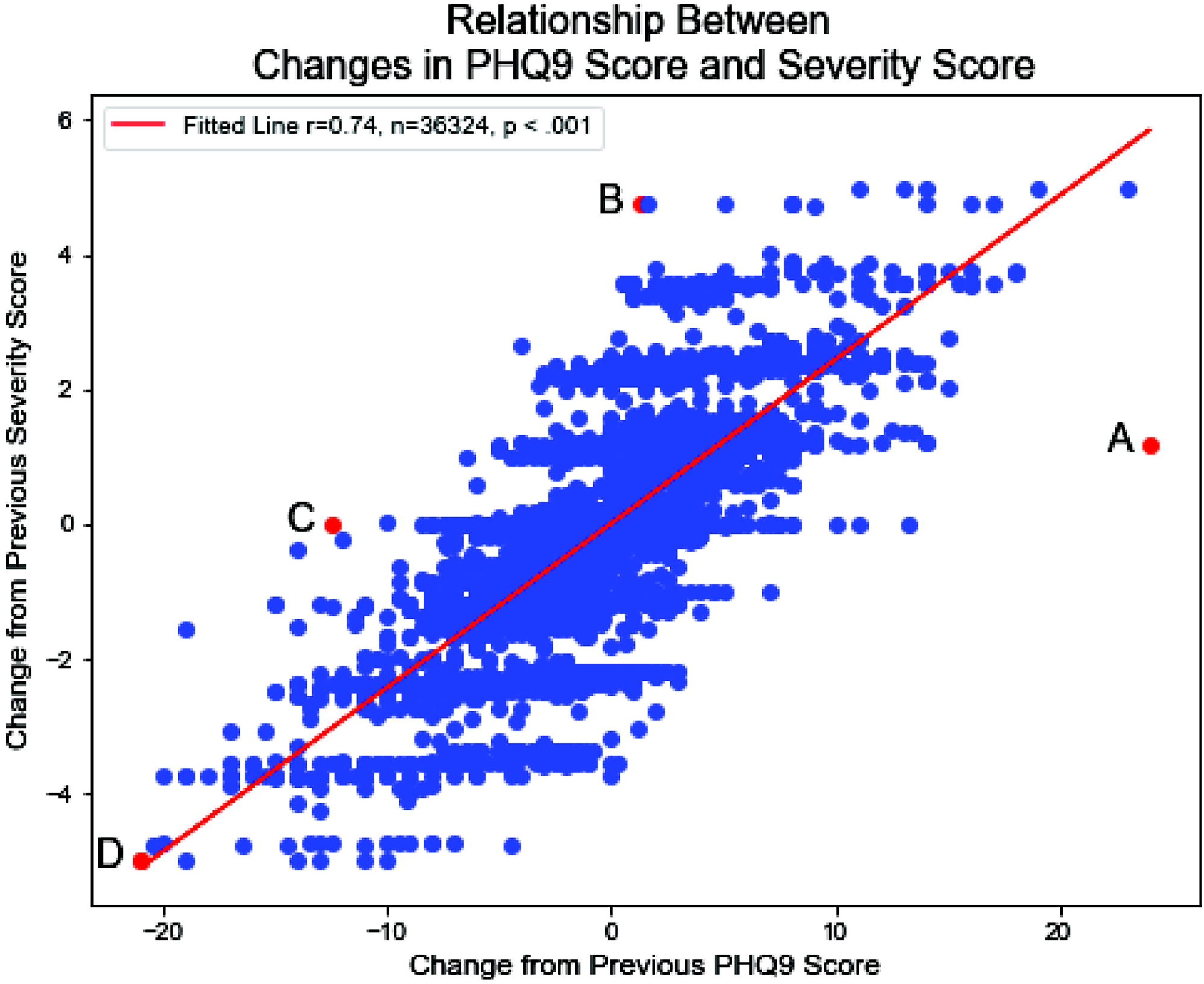
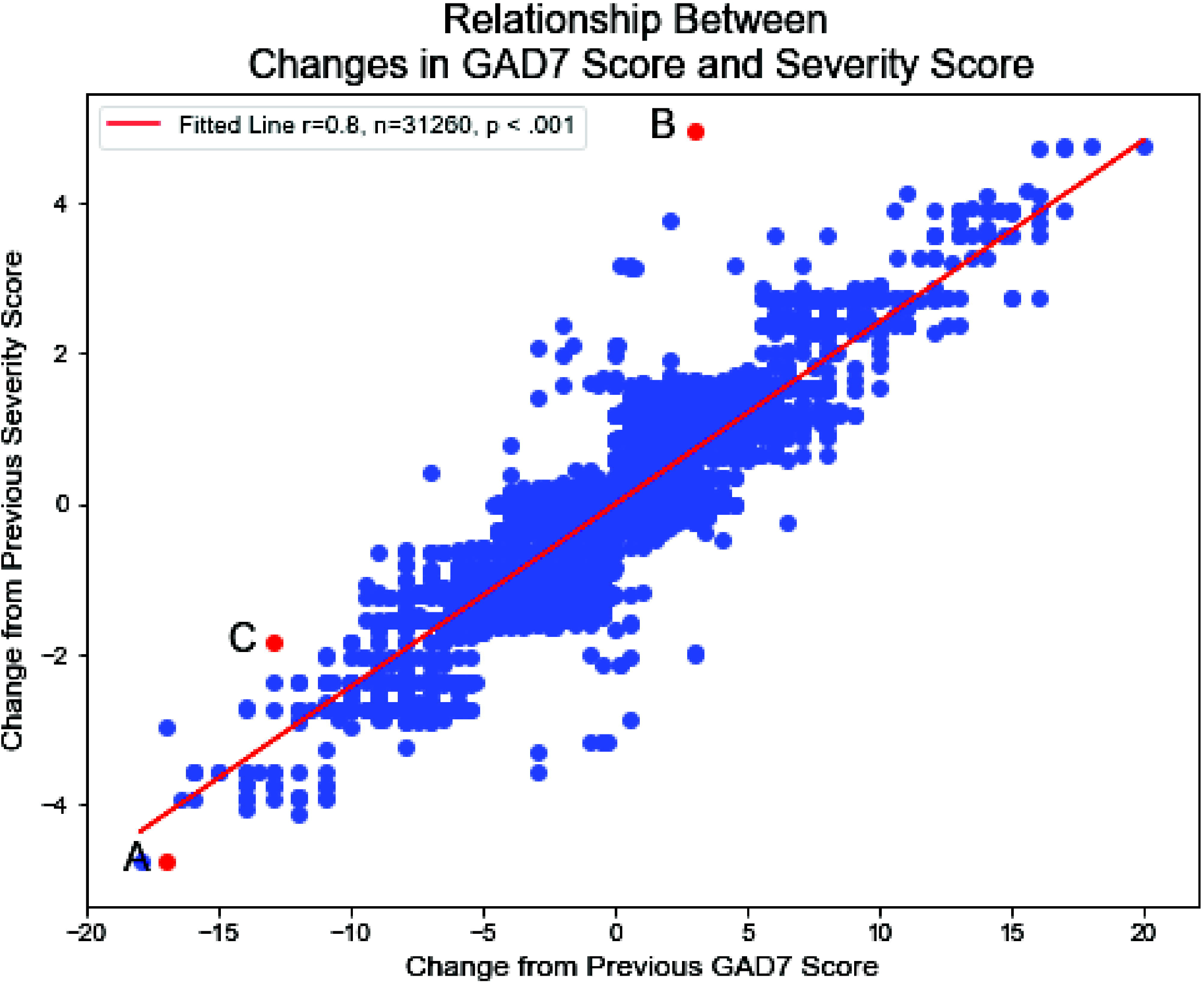
Overview of the process for developing and assessing the NeuroFlow Severity Score (NFSS).

### Comparisons to PHQ-9 and GAD-7

The PHQ-9 is a 9-item questionnaire used to screen for depression in medical settings. Each individual item is scored as “0” (not at all) to “3” (nearly every day) for a potential total score of 0 to 27. PHQ-9 scores of 5, 10, 15, and 20 represent mild, moderate, moderately severe, and severe depression, respectively. During the course of MBC, patients are asked by their care provider to complete these validated assessments, typically every 2 to 4 weeks. The clinical purpose of these assessments is to help support clinicians in making a diagnosis, to quantify depression symptoms, and to monitor changes over time to see if treatment is making a difference.^25,26^ The GAD-7 is a 7-item questionnaire used to screen for anxiety in medical settings; individuals are asked how often they have experienced certain feelings in the previous 2 weeks on a scale of 0 to 3 (not at all, several days, more than half the days, nearly every day). The total possible score is 21 and scores of 5, 10, and 15 are considered cut-offs for the presence of mild, moderate, and severe anxiety respectively.^27^ These are components of the measurements that patients complete in the NeuroFlow platform such that for every NFSS, there is a corresponding PHQ-9 and/or GAD-7 score for that individual at that point in time.

Using de-identified records, the NFSS for a given individual was plotted against their GAD-7 (n=31,260) and PHQ-9 (n=36,324) scores. For each comparison, we used linear regression to fit the slope of the line to our data and used Pearson’s product moment correlation coefficient (r) to quantify and summarize the direction and magnitude of the relationship between our variables. The Pearson product moment correlation coefficient, r, can be any number between -1 and 1 with the sign of r corresponding to the direction of the relationship between our variables and the number corresponding to the magnitude of the relationship between our variables. When r is positive, as one variable increases, the other increases as well. When r is negative, as one variable increases, the other decreases.

The NFSS scores and PHQ-9 scores were also compared using the Bland-Altman analysis, which compares the differences between 2 measures vs the mean of the 2 measures. This analysis evaluates whether or not 2 measurement methods return the same results and therefore could be used interchangeably.^30^ Because PHQ-9 scores vary from 0 to 27 and NFSS scores vary from 1 to 5, we first converted PHQ-9 scores to the clinically meaningful categories of 1 for no depression on PHQ-9 (score 0-4 on PHQ-9), 2 for mild depression (scores 5-9 on PHQ-9), 23 for moderate depression (scores 10-14 on PHQ-9), 4 for moderately severe depression (scores 15-19 on PHQ-9), and 5 for severe depression (scores 20-27 on PHQ-9). Differences between the converted PHQ-9 score and the NFSS were then plotted against the average of the converted PHQ-9 and the NFSS.

## Comparison to Clinical Assessments

A panel of 6 behavioral-health expert clinicians provided 2 clinical assessments for each of 250 individuals based on 2 different blinded presentations of data from de-identified patient records. As shown in Table 1, the first data set provided only 30-day average PHQ-9 and GAD-7 scores recorded in the app over a 30-day period for these 250 individuals. The second data set provided measures included in the NFSS in addition to the 30-day average and maximal PHQ-9 and GAD-7 scores, including the suicidal ideation score from the PHQ-9, measures of sleep and mood, whether the individual reported working with a behavioral health specialist, and whether severe depression or anxiety had been present in the prior 30-day period. The 2 datasets were randomized independently to ensure that the order of records was not repeated. Clinicians were asked to assign a clinical rating of symptom severity to each of the 250 individual records for both data sets, using a scale of 1 (low to minimal) to 5 (severe) (Table 2).

**Table 1.** Variables Included in Dataset 2 Evaluated by Expert Panel

| Variable | Possible score(s) | Description |
| --- | --- | --- |
| avg_phq9_score | 0-27 | The average phq-9 score. |
| max_phq9_score | 0-27 | The highest phq-9 score recorded. |
| q9_max | 0-3 | The highest score recorded for the 9th question of the phq-9, which screens for suicidal ideation as follows:<br>“Over the past two weeks, how often have you been bothered by thoughts that you would be better off dead or of hurting yourself in some way.<br>0=Not at all<br>1=Several days<br>2=More than half the days<br>3=Nearly every day" |
| avg_gad7_score | 0-21 | The average gad-7 score. |
| max_gad7_score | 0-21 | The highest gad-7 score recorded. |
| avg_sleep_rating | 0 (best)-4(worst) | The average sleep rating. |
| sleep_count | 0-30 | The number of sleep ratings. |
| avg_mood_rating | 0 (best)-4(worst) | The average mood rating. |
| mood_count | 0-30 | The number of mood ratings. |
| has_bh_specialist | 0=no; 1=yes | This value denotes whether or not a patient self-reported having a behavioral health clinician. |
| is_severe | 0=no; 1=yes | This value denotes whether a severe gad7 or phq9 score was recorded in the 30-day period before this period. |

NOTES: 1. All scores except "is\_severe" are for the same 30-day period; is\_severe is gathered from the prior 30-day period, 2) -1 denotes that the measure was not recorded in the 30-day period.

**Table 2.** Clinician symptom severity rating scale

|  |  |
| --- | --- |
| 1 | Low to minimal severity |
| 2 | Mild severity |
| 3 | Moderate severity |
| 4 | Moderately severe |
| 5 | Severe |

For each data set, the mean clinician expert score was plotted against the NFSS for the same individual (n=250). Linear regression was calculated, and both the Pearson correlation coefficient and the Kendall tau measure were used to evaluate the relationship between the mean clinician expert score and the NFSS. See Figure 2 for an overview of how the NFSS was compared to clinical assessments.

**Figure 2.**
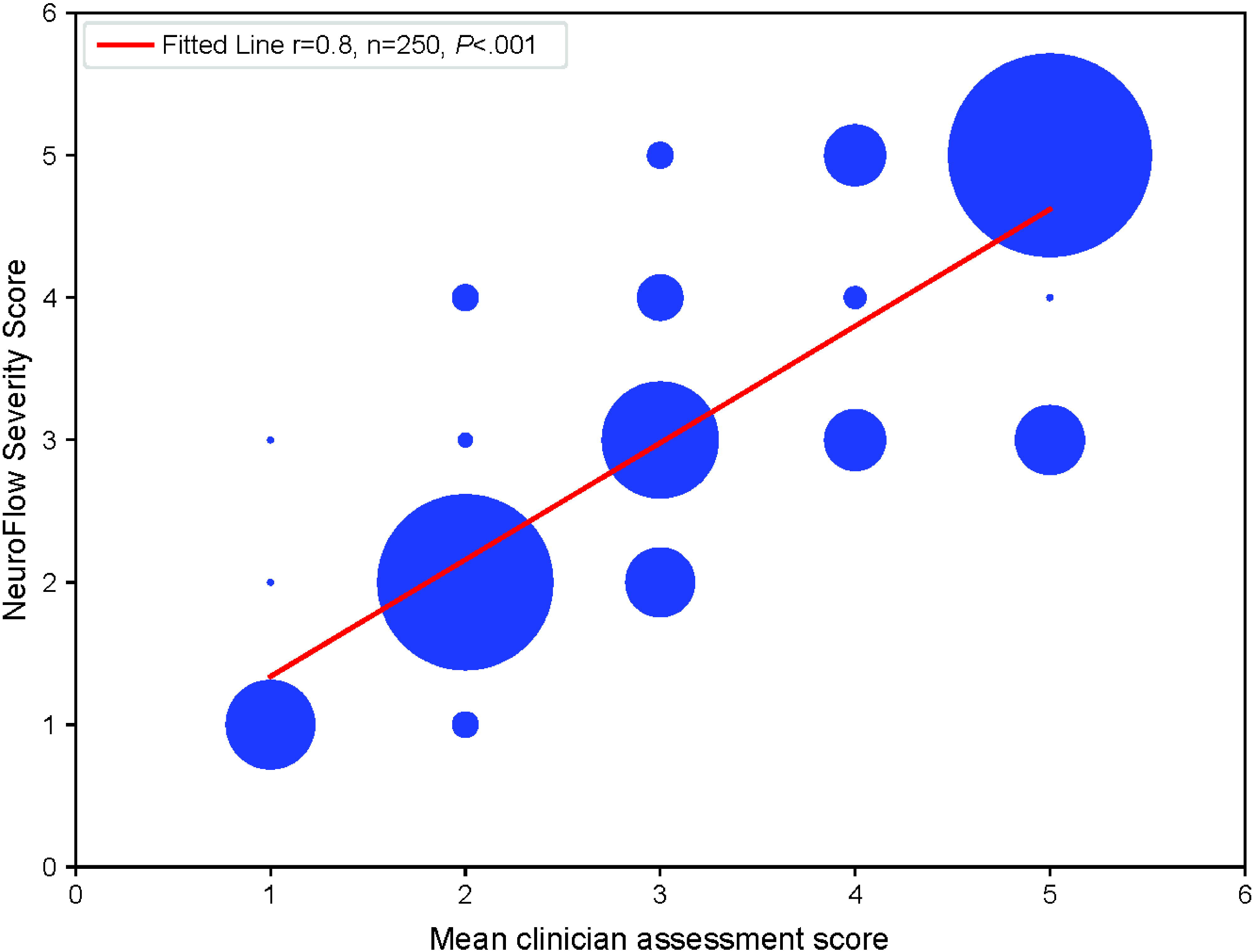
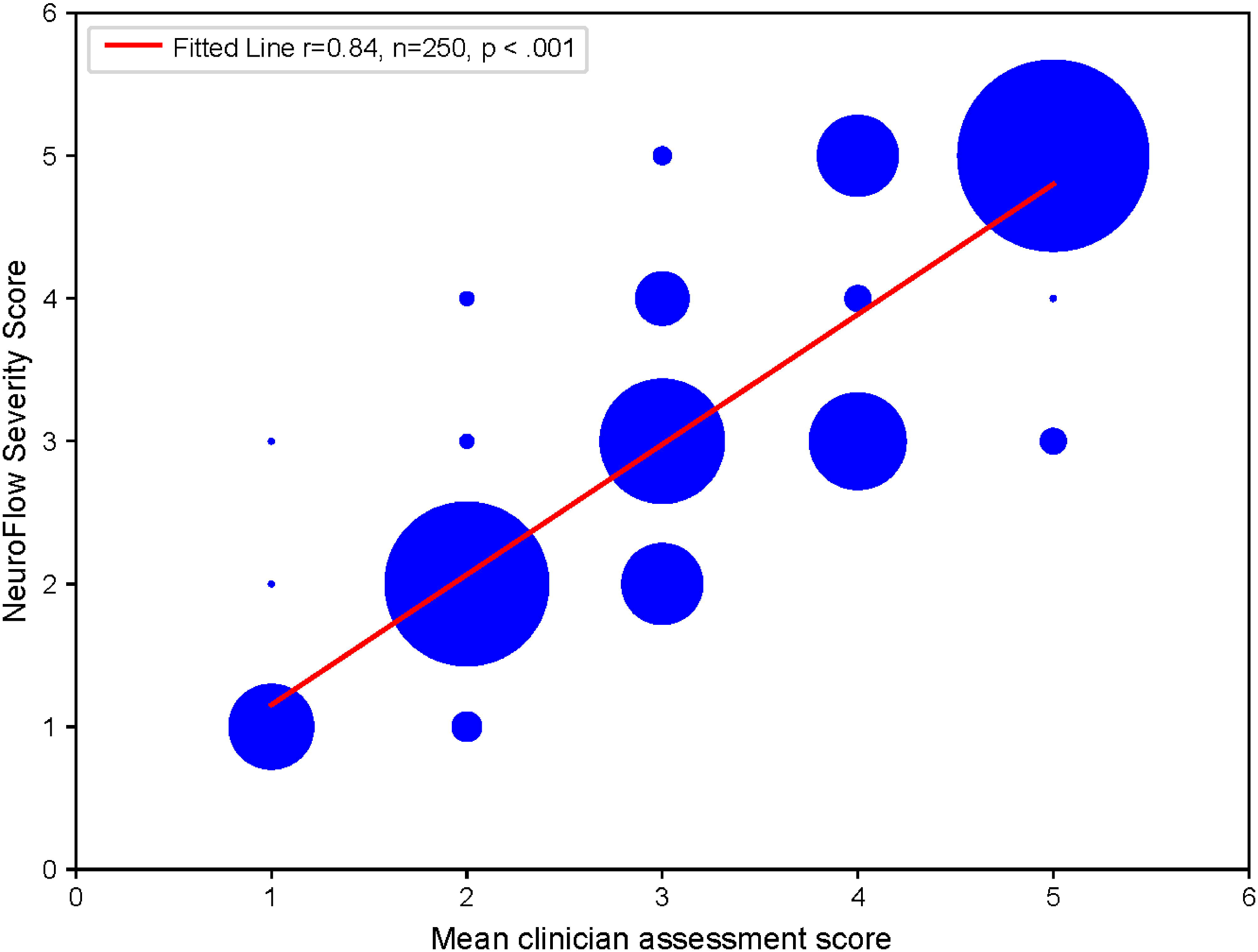
Overview of the process for comparing the NeuroFlow Severity Score (NFSS) to clinician’s assessments.

## Results

Normal distribution of data points around the mean and median were found for all 3 changes measured (ie, NFSS, PHQ-9, and GAD-7). The data for each measure were homoscedastic, supporting the use of linear regression for analysis of possible correlations.

After plotting the change in NFSS vs change in PHQ-9 scores for each individual record, we found a strong positive correlation of change in NFSS with change in PHQ-9 score (r=.74, P<.001; Figure 4). Similarly, changes in the NFSS were strongly and positively correlated with change in the GAD-7 score (r=.80, P<.001; Figure 4).

**Figure 3.**
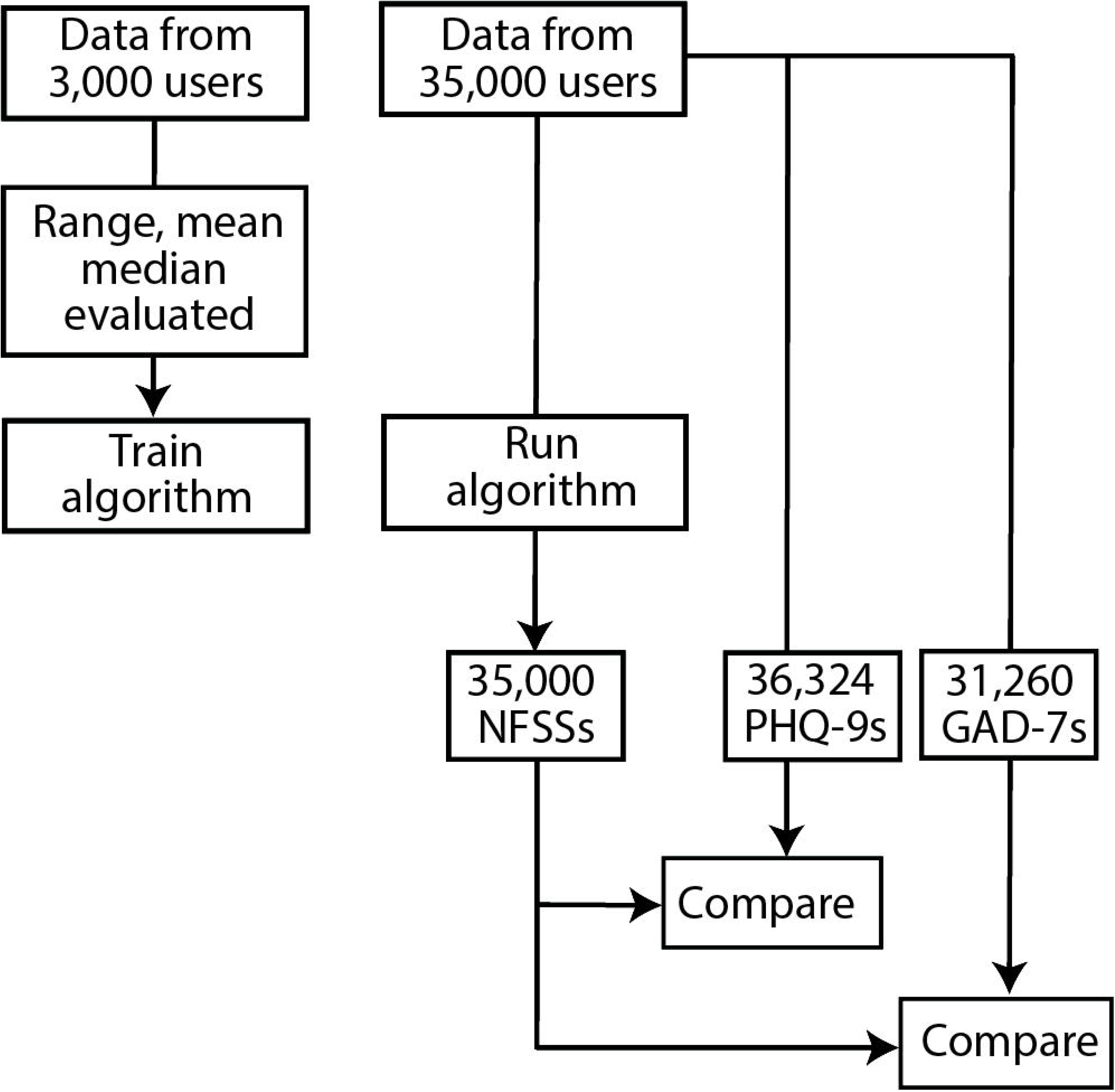
Bland Altman analysis shows that the PHQ-9 and NFSS are significantly different measures with differences increasing around the mean of the 2 measures.

**Figure 4.**
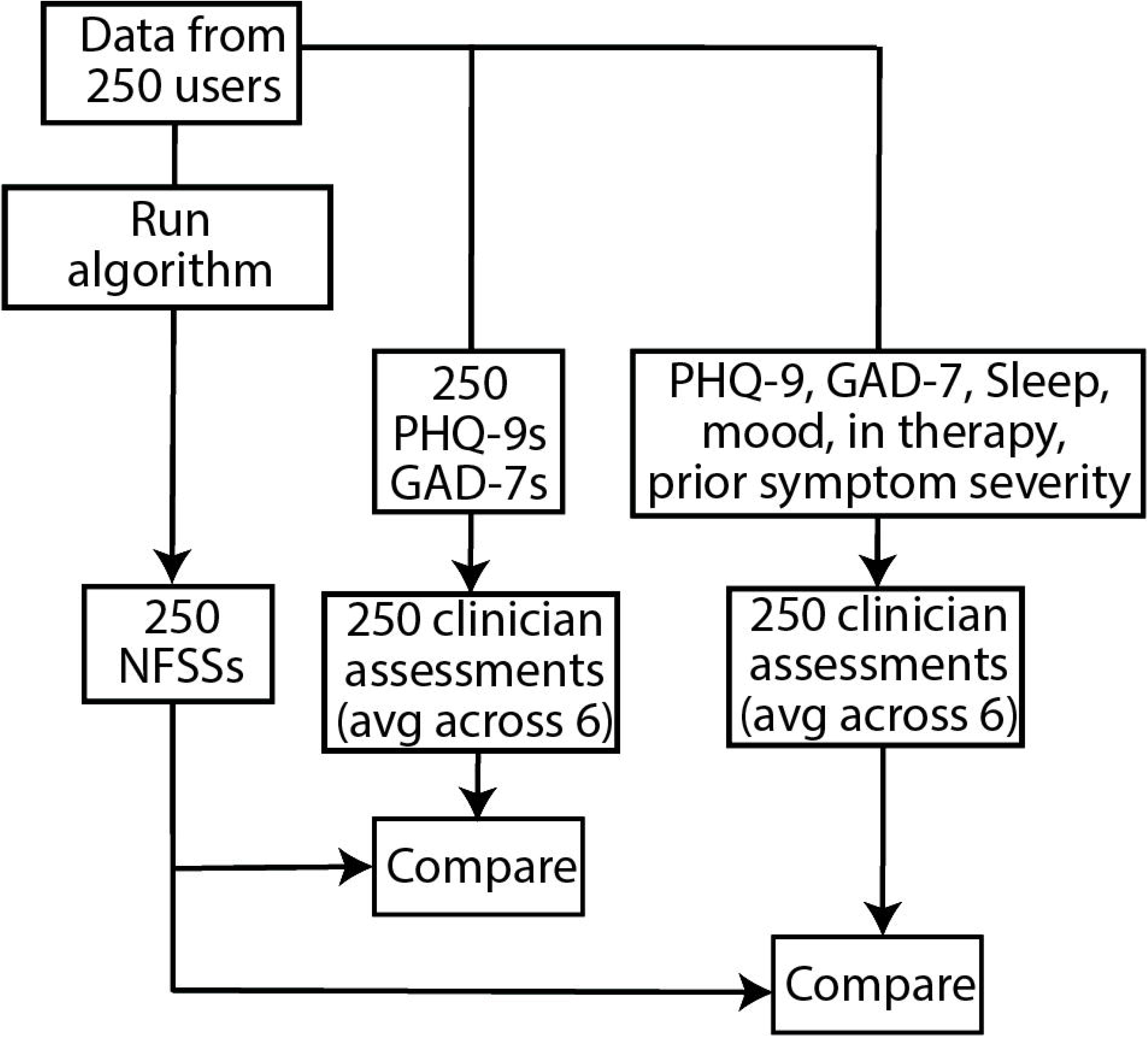
Change in NeuroFlow Severity Score (NFSS) vs change in Personalized Health Questionnaire-9 (PHQ-9) score (Left) or Generalized Anxiety Disorder-7 (GAD-7) over a 30-day period. Linear regression for NFSS vs PHQ-9 has a correlation coefficient (r) of .74 (n=36324; P<.001) and for NFSS vs GAD-7, r=.80 (n=31260, *P*<.001) showing a strong correlation between change in the NFSS and PHQ-9 and GAD-7 scores. Specific records (A-D) that appear to be outliers that could be caused by noise or artifact were confirmed as instead being instances when change in the NFSS but not PHQ-9 or GAD-7 scores correctly identified an individual’s level of risk for depression or anxiety based on expert clinician review.

We also evaluated several data points that did not fit the linear regression and found these were not errors or anomalies, but rather were cases in which the NFSS provided clinically meaningful information not captured by the PHQ-9 or GAD-7 score alone. For example, an individual whose PHQ-9 score changed from reflecting minimal to moderately severe symptoms of depression (Figure 4A, point A) had a smaller change in the NFSS because the NFSS score incorporated their active involvement in mental health treatment and absence of suicidal ideation. Another individual, in contrast, had PHQ-9 score changes that reflected a decrease in symptoms of depression but did not have a drop in the NFSS because the NFSS incorporated ongoing thoughts of self-harm/suicidal ideation reported by this individual.

On the Bland-Altman analysis comparing the PHQ-9 with the NFSS, we found that the NFSS measure is significantly different from the PHQ-9 with the difference between the 2 measures increasing around the mean of the 2 measures (Figure 3).

There were also strong correlations between the NFSS and clinical experts’ assessments both when the expert clinical score was generated from only average PHQ-9 and GAD-7 scores (Figure 5, r=.80, P<.001) and when the larger dataset was used by the clinicians (Figure 5, r=.84, P<.001).

**Figure 5.**
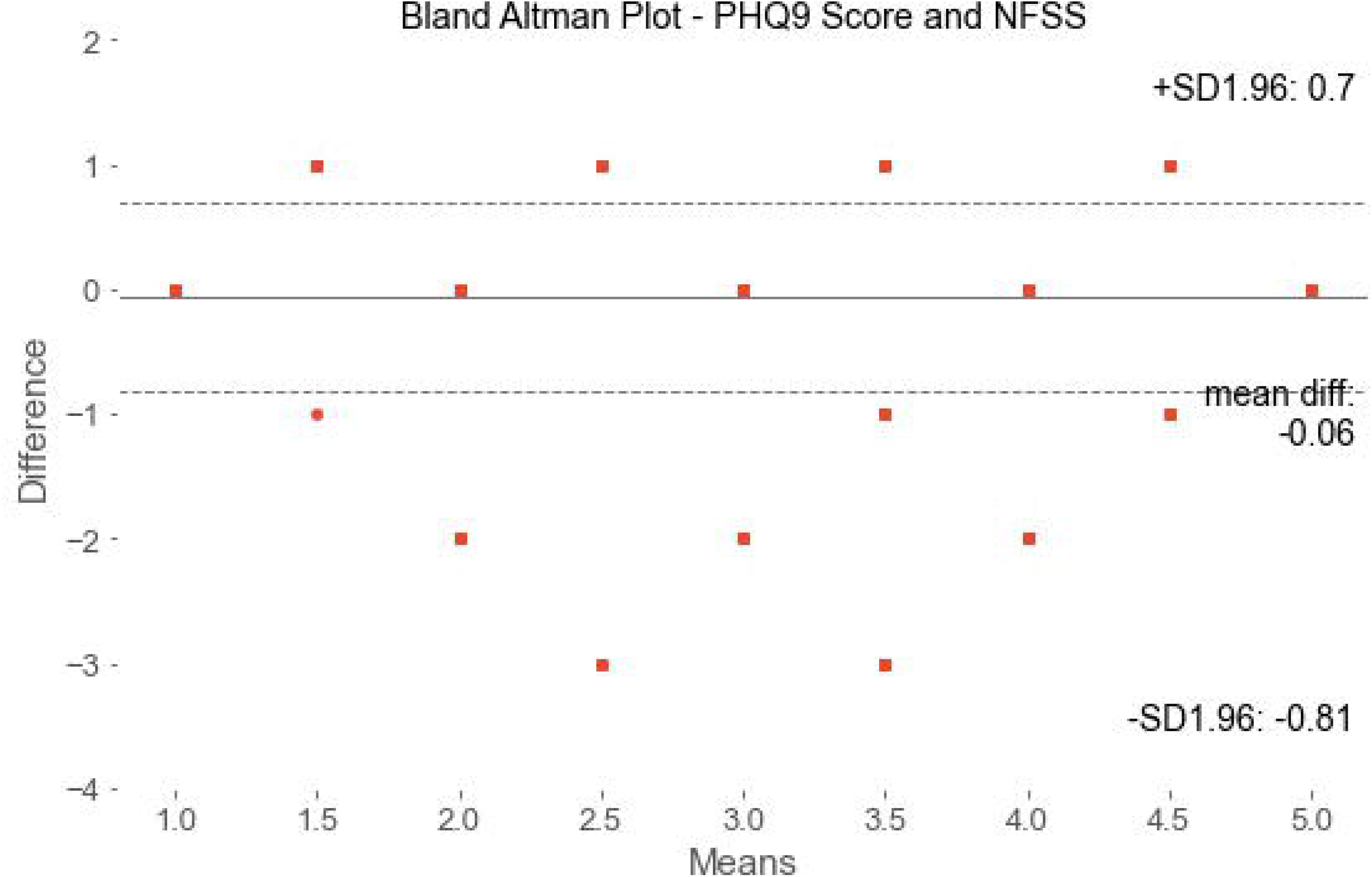
NeuroFlow Severity Score (NFSS) compared to mean clinical assessments score by expert clinician panel who reviewed 30-day averaged Personalized Health Questionnaire-9 (PHQ-9) score and Generalized Anxiety Disorder-7 (GAD-7) scores (Left) or 30-day PHQ-9 and GAD-7 average and maximum scores, sleep ratings, mood ratings, presence of suicidal ideation,and presence of severe symptoms in the prior month (Right). Linear regression analysis showed strong correlation between the NFSS and clinician assessments for both datasets, with a slightly stronger correlation for the more comprehensive data set (r=0.84, *P*<.001) vs just the average PHQ-9 and GAD-7 scores (r=.80, *P*<.001). The 2 datasets came from the same 250 individuals and were separately randomized to ensure records were presented in different orders.

## Discussion

Evidence-based treatment of depression and anxiety remains an unmet need contributing to a large burden of disease and cost to society of over $210 billion annually for depression alone.^24^Despite strong evidence that MBC improves outcomes, fewer than 10% of people with depression or anxiety are able to access such care.^14,15^ Integrating MBC into primary care has been proposed as a means to address the shortage and unequal geographic distribution of behavioral health specialists that contributes to this undertreatment.^17,19,29^

In practice, integrating MBC into primary care has proven effective, but is still not widely adopted.^16^Simply screening for the presence of depression or anxiety was done in only 4.3% of primary care visits in 2016, for example.^19^ Barriers to implementation of MBC in integrated behavioral health include systemic factors such as the lack of a consistently used measurement system and resources for training staff to use a measurement system in a HIPAA-compliant manner. In behavioral health care, HIPAA compliance is of particular importance because of the stigma around behavioral health disorders that leads to discrimination and decreased quality of life.^5,6,12-15,30^ On an individual level, concerns about confidentiality and the time spent taking measurements are major barriers to utilization of MBC. In addition, finding time for measurement of behavioral health symptoms during primary care visits--already considered too short for the number of clinical tasks that must be done--remains a challenge. The NeuroFlow platform and the NFSS were developed with a goal of facilitating MBC integration into primary care with decreased burden by taking the measurement aspect outside of the clinical appointment and into the patients’ daily activities with an engaging smartphone or desktop application.

Here we describe the development of a new measure for clinically significant depression and anxiety, the NFSS. The NFSS is derived via an artificial-intelligence powered algorithm that uses both patient-reported measures taken outside the clinical visit and passively collected data through use of a digital application. Regression analysis shows that the NFSS correlates strongly with both the PHQ-9 score and the GAD-7 score, which are commonly used validated measurement scales for depression and anxiety with .88 sensitivity and specificity for the PHQ-9 and .83 sensitivity and .84 specificity for the GAD-7.^25-27^ Additionally, the NFSS correlates strongly with clinician assessment based on PHQ-9/GAD-7 scores alone (r=.80, P<.001) or on analysis of larger number of data reported via the Neuroflow platform (r=.84, P<.001). These strong correlations suggest that the NFSS can be used as a proxy measure for the presence of depression and anxiety. If, for example, a clinician saw at a glance (on the clinician dashboard or EHR) that a patient had an NFSS of 2 or more, they would know the need to address depression or anxiety was present and could even schedule a separate follow up visit for these concerns as needed. Although the expert clinician panel’s assessments in this study were made with review of recorded data measures rather than in-person clinical assessments, the strong correlations with both clinician assessment and validated measures provide further confidence in the NFSS for clinical use.

At the same time, Bland-Altman analysis shows that the NFSS is significantly different from the PHQ-9, which likely reflects the larger number of measures and the use of weighting for more or less significance for depression or anxiety based upon comparison to a large set of individuals. Analysis of outliers in regression analysis showed instances in which the NFSS more accurately identified people with or without clinically significant depression or anxiety than the PHQ-9 or GAD-7 scores did, respectively. The identification of the NFSS as a measure that is different and still correlates with both validated scales and clinician assessment suggests it can be used as a proxy for those validated measures and also that it may have greater utility than those measures alone.

Although the PHQ-9 and GAD-7 combined comprise only 16 questions, administering these measures during the primary care visit can be challenging, in part because of time constraints. To add just 1 of these 2 measures, for example, could take 20% to 40% of an already short 17 to 21 minute long primary care visit. Rather than shortening the time taken for measurement, the NeuroFlow platform aids in MBC integration by taking measurement out of the clinical visit and into the day-to-day life of the patient. Using both data reported by patients through use of the app and passively collected data, the NFSS provides a score that is inclusive not only of the full PHQ-9 and GAD-7 data but also other meaningful measures. By assessing a larger number of parameters with an artificial intelligence-driven algorithm, the NFSS may be able to identify at-risk individuals who may have been missed with just a PHQ-9 or GAD-7 assessment. In addition, digital delivery of the NFSS in the patient’s EHR makes it such that the screening is HIPAA-compliant, addressing any confidentiality concerns, as well as already completed when the patient is with the clinician. This may allow the clinician to use the minutes that would have been taken for screening to instead use that time for further assessment and suggestions for treatment, as appropriate.

Providing a system-wide, HIPAA-compliant measurement system that requires minimal to no training of medical practice personnel, removes any paper-and-pencil assessments and digitization/storage needs, and integrates measurement not only into primary care but also into the day-to-day life of individual patients addresses many of these barriers. From the perspective of clinicians, the NFSS is valuable in that it provides a single score of 1 to 5 that is generated from daily day-to-day platform use by the patient, can be integrated into the EHR, can alert the clinician to the presence of concerning endorsements of depression and anxiety symptoms, which in turn can prompt further assessment, treatment, and referrals as needed.

Another advantage of the NFSS is that it seamlessly incorporates measurement of behavioral health symptoms into a person’s typical day-to-day activities that includes use of their mobile phone. Taking a moment (∼30 seconds) to track mood or sleep for multiple days of the week is less onerous for the patient compared with the 4-8 minutes needed to take both the PHQ-9 and GAD-7.

As with all health-related apps, there is appropriate concern for whether or not individuals will use the app and record data consistently. In this regard, the NFSS has 2 particular advantages. First, the platform has a high rate of user retention at 70%, which is 21 times higher than the typical 30-day retention of 3.3% for health-related apps.^24^ Second, incorporation of additional patient reported measures (e.g., sleep rating, mood rating, number of times a measure was tracked) in addition to PHQ-9 and GAD-7 scores means that a reliable severity measure is available even if 1 particular measure (eg, the PHQ-9 or GAD-7 score) is not present for a particular individual.

## Conclusions

The NFSS is a digitally-derived measurement of symptoms of depression and anxiety generated with an artificial-intelligence powered algorithm developed from analysis of 3,000 Neuroflow users. Comparison of over 35,000 NFSSs with PHQ-9 and GAD-7 scores taken at the same point in time by the same individuals showed a strong correlation between the NFSS and both the PHQ-9 and GAD-7, respectively. Clinician assessment of PHQ-9 and GAD-7 scores also correlates strongly with the NFSS. Together, these correlations strongly suggest that the NFSS can be used as a proxy measure for the presence of depression and anxiety. Bland-Altman analysis, however, shows that the NFSS is a significantly different measure. Prospective studies to further measure the sensitivity, specificity, and clinical noninferiority of the NFSS are thus warranted.

## Data Availability

Deidentified data are available from the corresponding author for researchers who meet the criteria for access and collaboration.

## Notes

### Competing Interest Statement

Authors Adam Pardes and William Lynch are employees of NeuroFlow, Inc. Author Michael L. Platt is an
advisor to NeuroFlow, Inc.

### Funding Statement

No external funding was received for this study.

### Author Declarations

Exemption provided by Advarra IRB (ID: Pro00046380)

